# A mitoxyperilysis-derived spatial signature predicts immune exclusion and survival in oral squamous cell carcinoma

**DOI:** 10.64898/2026.09.04.26362241

**Authors:** Jianlin Liu, Dongyan Zhang, Zhen Meng, Bo Zou

## Abstract

Immune checkpoint blockade offers limited clinical benefit to patients with oral squamous cell carcinoma (OSCC) harboring immune-excluded microenvironments. The spatial mechanisms maintaining this stromal barricade remain unresolved. Analyzing single-cell and spatial transcriptomics across multiple cohorts, we identified a localized immunosuppressive network driven by mitoxyperilysis—a mitochondria-dependent lytic cell death process. Quantitative spatial mapping revealed that ISG+ PMN-MDSCs and GZMB+ pDCs accumulate at the tumor invasive margin, where they establish a physical and metabolic barrier restricting cytotoxic T-cell infiltration. Computational network ablation confirmed that both subsets are required to maintain this exclusion. We then derived a 5-gene prognostic signature (*CTSG, NAMPT, AREG, PLAU, CXCL8*) directly from this intercellular communication network. Across independent cohorts, this signature captured an immune desert phenotype and independently predicted patient survival, further improving risk stratification when integrated with standard clinical staging. Our findings define a structural immune evasion axis in OSCC, offering a practical tool to guide combination therapies aimed at remodeling the myeloid-stromal barrier.

## 1. Introduction

Oral squamous cell carcinoma (OSCC) represents a heterogeneous malignancy characterized by a highly immunosuppressive tumor microenvironment (TME) ^1–3^. Although immune checkpoint blockade therapy (ICB) offers a new therapeutic option^4^, clinical benefits appear to be confined to a minority of patients ^5^. While the PD-L1 combined positive score (CPS) is currently utilized to predict therapeutic response, additional biomarker stratification remains essential to inform the value-based integration of immunotherapy among patients with OSCC^6^.

Mechanistically, it is widely accepted that ICB therapy exerts a more profound effect on “;hot” tumors (immune-inflamed phenotype), while demonstrating limited efficacy against “;cold” tumors (immune-desert phenotype) or “;altered” tumors (immune-excluded phenotype) ^7–9^. Combination therapies are required to enhance immune cell infiltration into tumor tissue and convert “;cold” or “;altered” tumors into “;hot” ones. In OSCC, this non-responsive “;cold” or “;altered” state is shaped by a spatial organization, wherein cytotoxic T cells are physically or biochemically restricted from infiltrating the malignant parenchyma, accumulating instead in the surrounding stroma^10^. However, the mechanisms establishing these physical barriers remain incompletely understood.

Emerging evidence indicates that localized metabolic stress can remodel the structural organization of the TME^11^ ^12^. Recently, mitoxyperilysis was characterized as a distinct mitochondria-dependent lytic cell death mechanism, which is induced by metabolic disruption and innate immune signaling, and driven by localized mitochondrial oxidative damage^13^. Analogous to other lytic death modalities such as pyroptosis, which remodels the tumor immune microenvironment by releasing damage-associated molecular patterns (DAMPs) and pro-inflammatory mediators to recruit immunosuppressive cells (e.g., MDSCs and Tregs), mitoxyperilysis is poised to drive immune evasion^14^. However, the cellular lineages executing this terminal stress response and their role in shaping the OSCC immune-excluded landscape are currently unknown.

In this study, we integrated single-cell and spatial transcriptomic atlases to characterize the physical architecture of the OSCC immunosuppressive barrier. We identified a localized intercellular crosstalk governed by two specific innate immune subsets_—_ISG+ PMN-MDSCs and GZMB+ pDCs_—_exhibiting mitoxyperilysis-associated transcriptional signatures at the invasive margin. By spatially resolving these two subsets and their communication networks, we mapped the spatial exclusion of cytotoxic T cells within OSCC. To translate these spatial insights into clinical utility, we utilized LASSO Cox regression to identify a 5-gene prognostic signature (*CTSG*, *NAMPT*, *AREG*, *PLAU*, and *CXCL8*). This provides a validated clinical tool that bridges high-resolution TME biology with patient risk stratification.

## 2. Materials and Methods

### 2.1 Data acquisition

Matched single-cell RNA sequencing (scRNA-seq) and spatial transcriptomic (ST) data for oral squamous cell carcinoma (OSCC) were obtained from the Gene Expression Omnibus (GEO) under accession number GSE310797 ^15^. This cohort comprises 16 OSCC samples profiled using the Mobinova-100 system, with a subset of six matched samples processed via the 10x Genomics Visium spatial platform. For prognostic modeling, we downloaded bulk RNA sequencing data and corresponding clinicopathological profiles of head and neck squamous cell carcinoma (HNSCC) patients from The Cancer Genome Atlas (TCGA-HNSC), and obtained an OSCC-specific cohort through strict anatomical site selection ^16^. Raw count matrices were then log2-transformed and normalized as Counts Per Million (log_2_(CPM + 1)) to correct for sequencing depth biases and stabilize data variance for downstream linear modeling. An independent bulk transcriptomic cohort (GSE41613) ^17^ was acquired from GEO for external validation.

### 2.2 Single-cell RNA-seq data processing and subset annotation

Single-cell RNA-seq data were processed using the Seurat R package (Version 5.1.0) ^18^. To ensure high data quality while capturing metabolically stressed cells intrinsic to the TME, we rigorously filtered out low-quality cells, retaining those with >200 expressed genes (nFeature_RNA > 200) and a mitochondrial expression fraction below 20% (percent.mt < 20%). Following log-normalization (*NormalizeData*) and the identification of highly variable genes (*FindVariableFeatures*), individual samples were directly merged to preserve the authentic patient-specific metabolic microenvironments without artificial over-correction.

Dimensionality reduction and visualization were performed using PCA and UMAP. Following initial unsupervised clustering and broad lineage annotation via canonical markers, major immune compartments of interest were subjected to sub-clustering. Finally, differentially expressed genes (DEGs) defining these fine-resolution subsets were computationally identified.

### 2.3 Mitoxyperilysis signature quantification and trajectory inference

A 7-gene mitoxyperilysis signature ^13^, comprising the mTORC2 complex (RICTOR, MTOR, MLST8, MAPKAP1) and the apoptosis execution axis (BAX, BAK1, BID), was quantified using the *AddModuleScore* function. To infer biological pathways, the identified DEGs were mapped across the GO and KEGG databases via the clusterProfiler package (Version 4.12.6) ^19^. Following the computational exclusion of non-myeloid contaminants, the developmental continuum of the myeloid compartment was modeled using Monocle 3 (Version 1.3.7) ^20^, with the trajectory root biologically anchored to the monocyte-enriched state.

### 2.4 Cell-cell communication and in-silico perturbation analysis

Cell-cell communication networks and their global topologies were constructed using the *CellChat* R package (Version 2.1.2) and its *netAnalysis_computeCentrality* function^21^. To computationally ablate the target cell populations, an *in silico* matrix perturbation approach was employed, forcing all their associated incoming and outgoing signaling probabilities to zero. The resulting absolute flux loss (ΔFlux) within the remaining network was then measured against the baseline condition. Finally, co-regulated signaling modules were resolved through hierarchical clustering coupled with K-means partitioning and visualized with the *ComplexHeatmap* package (Version 2.20.0) ^22^.

### 2.5 Spatial transcriptomics (ST) mapping and geometric engagement modeling

By calculating the geometric mean of library size-normalized ligand and receptor expression across spots, we established a Spatial Interaction Density Index (SIDI) to unbiasedly identify the optimal tissue slice exhibiting the highest signaling density for downstream intercellular modeling. Spatial signatures for target cell subsets were mapped via the *AddModuleScore* function using their top 20 respective marker genes. To computationally delineate the malignant epithelial boundary, a 2D Gaussian kernel density estimation was applied based on the differential tumor-versus-stroma score (*S*_Δ_), mapping strictly to the intrinsic spatial coordinates derived from the initial histological alignment.

To quantify localized immunosuppressive interactions, a geometric engagement index (*E_i_*) was developed for each spatial spot (*i*):

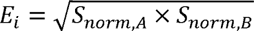

where *S_norm_* represents the min-max normalized signature score of the target populations. The geometric mean was applied to impose a biological “;AND” logic gate, ensuring that high engagement scores require the spatial co-enrichment of both interacting components, thereby penalizing asymmetric, mono-population background noise. Finally, a spatial masking algorithm restricted signaling hotzones to the physical boundaries of the target subsets.

### 2.6 Construction and validation of the prognostic signature

Patients with fewer than 30 days of follow-up were excluded to minimize non-tumor-specific confounding. Genes derived from the downstream intercellular communication networks were subjected to Least Absolute Shrinkage and Selection Operator (LASSO) Cox regression (glmnet v4.1.8) ^23^. The optimal penalty parameter (*λ*) was tuned via 10-fold cross-validation at the minimum partial likelihood deviance. Risk scores were computed by summing the coefficient-weighted expression values of retained genes. Patients were stratified by the median risk score for Kaplan-Meier survival analysis, and time-dependent ROC curves (timeROC ^24^) were generated to evaluate longitudinal predictive accuracy. The established model was externally validated using the GSE41613 cohort.

### 2.7 Immune microenvironment evaluation and spatial mapping

Single-sample Gene Set Enrichment Analysis (ssGSEA) was performed via the *GSVA* package (Version 1.52.3) ^25^ to quantify relative cell infiltration, and the expression disparities of core immune checkpoints between the two risk groups were evaluated. For clinical benchmarking, the signature’s predictive accuracy was compared against the established 18-gene Tumor Inflammation Signature (TIS)^26^ utilizing C-index. Finally, mathematical projection of continuous risk scores onto OSCC spatial coordinates allowed for the visual delineation of malignant boundaries and immune hotzones.

### 2.8 Nomogram construction and decision curve analysis

To facilitate clinical application, patient clinical stages were stratified into early (I-II) and advanced (III-IV) categories. A prognostic nomogram was subsequently constructed via multivariable Cox regression, integrating the signature-derived risk score with age and the stratified clinical stage. For internal validation, calibration curves assessing 1-, 3-, and 5-year overall survival were generated utilizing 1,000 bootstrap resamples. Decision Curve Analysis (DCA) ^27^ was performed to quantify the net clinical benefit across varying threshold probabilities, benchmarking the composite nomogram against either the risk score or clinical stage alone, implemented via the *rms* (version 8.1.0) and *survival* (version 3.8.6) packages.

### 2.9 Statistical Analysis

All analyses and visualizations were performed in R software (Version 4.4.1). Non-normally distributed continuous variables (e.g., expression levels, module scores) were compared using the Wilcoxon rank-sum test (two groups) or the Kruskal-Wallis test (multiple groups). Survival outcomes were evaluated via the Kaplan-Meier method with log-rank tests, followed by univariate and multivariable Cox proportional hazards regression to identify independent prognostic factors. Differences in model predictive accuracy (C-index) were assessed using the Z-test. The Benjamini-Hochberg procedure was applied to control the false discovery rate (FDR) during multiple hypothesis testing. Statistical significance was defined as a two-sided *P* < 0.05.

## 3. Results

### 3.1 Single-cell transcriptomic landscape identifies ISG+ PMN-MDSCs and GZMB+ pDCs as the cellular hubs of mitoxyperilysis

Unsupervised clustering of the OSCC single-cell transcriptomic atlas resolved 10 major cell lineages (Figure 1A), whose relative proportions exhibited inter-sample heterogeneity. Rather than being uniformly distributed, the mitoxyperilysis signature was selectively enriched across subpopulations, with B cells, CD8+ T cells, and myeloid lineages exhibiting significantly higher scores than the malignant epithelium (Figures 1B and 1C; *P* < 0.0001). Dissecting the signature components revealed lineage-specific expression profiles (Supplementary Figure S1). While CD8+ T cells expressed the upstream sensors (MTOR, RICTOR) and BAK1, they lacked the downstream effectors BAX and BID. Conversely, myeloid and B cells co-expressed BAX, BID, and MLST8; therefore, these two lineages were prioritized for sub-clustering.

**Figure 1.**
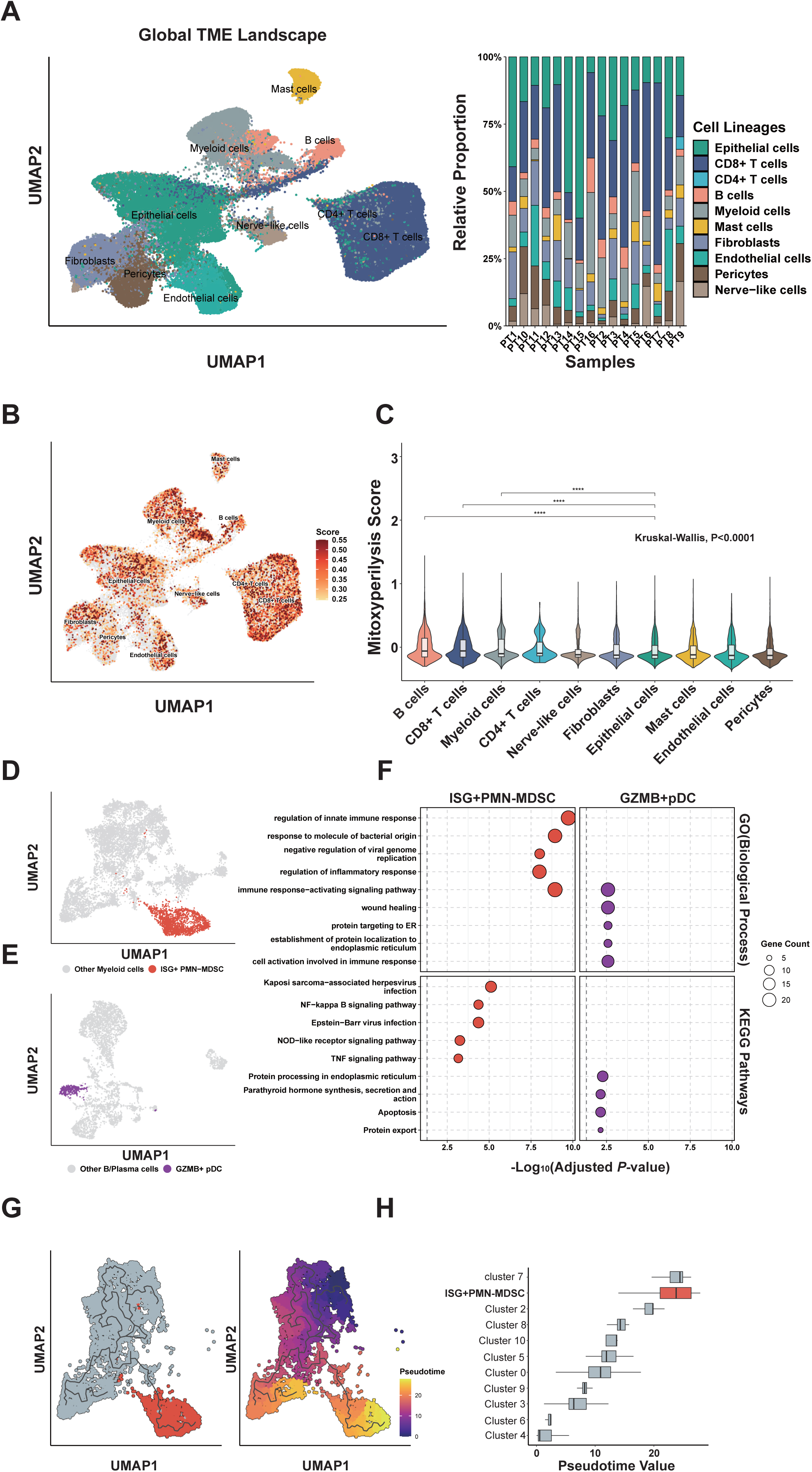
Single-cell transcriptomic landscape and enrichment of the mitoxyperilysis signature in OSCC. **(A)** UMAP visualization of the OSCC single-cell transcriptomic atlas identifying 10 major cell lineages. The bar plot illustrates relative proportions across samples. **(B)** UMAP visualization of mitoxyperilysis scores. The top 10% highest-scoring cells are highlighted. **(C)** Violin plots of mitoxyperilysis scores across major cell lineages. Significance was determined by the Kruskal-Wallis test, followed by Wilcoxon rank-sum tests for pairwise comparisons against malignant epithelial cells (P < 0.0001). **(D)** UMAP of the myeloid compartment, highlighting the ISG+ PMN-MDSC subset (Cluster 1, red). **(E)** UMAP of the B/plasma cell compartment, highlighting the GZMB+ pDC subset (Cluster 6, purple). **(F)** Faceted dot plot showing the top enriched GO biological processes and KEGG pathways for ISG+ PMN-MDSCs (left) and GZMB+ pDCs (right). Bubble size indicates gene count; color gradient represents negative log10-transformed adjusted P-values. **(G)** Pseudotime trajectory of the myeloid compartment originating from progenitor monocytes. **(H)** Boxplot of pseudotime distribution across myeloid subclusters.

Ranking the sub-populations by mean mitoxyperilysis scores identified Myeloid Cluster 1 (C1) and B-cell Clusters 6 (C6) and 11 (C11) as the top-scoring subsets (Supplementary Figure S2A, B). C11 was excluded due to limited cell numbers and the absence of specific effector markers (Supplementary Figure S2C, D and Supplementary Table S1). Although initially grouped within the B-cell lineage, C6 selectively expressed *LILRA4*, *CLEC4C*, and *GZMB*, identifying this cluster as GZMB+ plasmacytoid dendritic cells (pDCs) ^28^ ^29^. High expression of *ISG15* and *CXCL10* defined Myeloid C1 as ISG+ PMN-MDSCs ^30–32^. UMAP mapping visualized the distribution of these two distinct subpopulations (Figure 1D, 1E).

Gene Ontology (GO) and KEGG analyses further defined the transcriptomic profiles of these two subsets (Figure 1F). ISG+ PMN-MDSCs showed significant enrichment in the regulation of innate immune and inflammatory responses, as well as classical pro-inflammatory cascades (NF-kappa B, TNF, and NOD-like receptor pathways), While GZMB+ pDCs were enriched in pathways related to protein targeting to the endoplasmic reticulum (ER) and apoptosis signaling. Pseudotime trajectory inference revealed a pathway originating from monocytic states (Figure 1G). Although both Cluster 7 and the ISG+ PMN-MDSCs localized to the terminal stages of this developmental axis (Figure 1H), only the ISG+ PMN-MDSCs exhibited the elevated mitoxyperilysis signature scores.

### 3.2 Cell-cell communication networks reveal a hierarchical communication structure centered on GZMB+ pDC sinks and ISG+ PMN-MDSC emitters

Global network analysis identified GZMB+ pDCs as a dominant signaling sink with high incoming interaction strength (Figure 2A). Based on the weighted network structure (Figure 2B), malignant epithelial and fibroblast compartments acted as primary sources, funneling their strongest signals toward this pDC node. In contrast, ISG+ PMN-MDSCs served as signal emitters; they targeted the endothelial network and themselves, establishing paracrine and autocrine loops. Clustering the relative contribution matrices (Figure 2C) revealed that GZMB+ pDCs functioned as the principal receivers for the MIF and TGF-β signaling axes, while emitting TGF-β and EGF signals. ISG+ PMN-MDSCs served as the major receivers of the PLAU, CTSG, ANNEXIN, and IL1 pathways, while secreting PLAU, VISFATIN, CXCL, IL1, and OSM. The concurrent emission and reception of PLAU and IL1 by these MDSCs validated the autocrine loop operating within this compartment.

**Figure 2.**
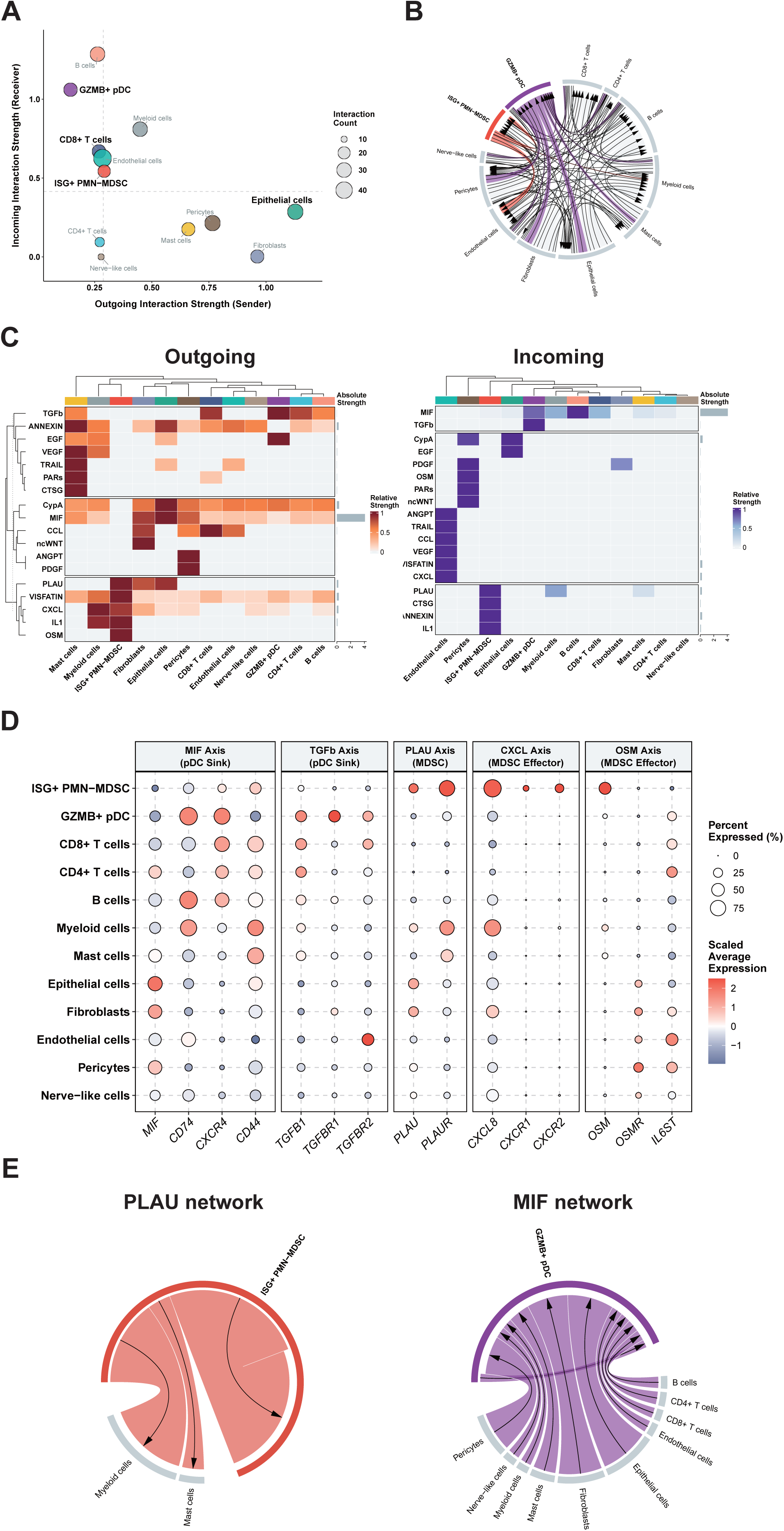
Cell-cell communication networks reveal a hierarchical structure centered on GZMB+ pDCs and ISG+ PMN-MDSCs. (A) Scatter plot of secreted signaling roles for all identified cell populations. The x- and y-axes represent outgoing and incoming interaction strengths, respectively. (B) Circle plot of the overall communication network. Edge width is proportional to the inferred interaction weight (probability sum), and node size reflects total signaling capacity. (C) Heatmaps of relative outgoing (left) and incoming (right) signaling patterns. Pathways are ordered via hierarchical clustering and partitioned by K-means (k=3). Right annotations display the absolute signaling strength of each pathway. (D) Faceted dot plot showing the expression of key ligands and receptors. Dot size represents the percentage of cells expressing the gene, and color intensity indicates the scaled average expression level. (E) Chord diagrams illustrating source-target interactions of core signaling axes. The left panel shows the outgoing PLAU network from ISG+ PMN-MDSCs, while the right panel highlights the incoming MIF network converging on GZMB+ pDCs.

Transcript-level evaluation of core ligand-receptor pairs corroborated these network inferences (Figure 2D). GZMB+ pDCs expressed the MIF receptors *CD74* (98.5%) and *CXCR4* (81.1%), alongside TGF-β receptors. ISG+ PMN-MDSCs expressed *CXCL8* (95.0%) and the inflammatory cytokine *OSM*. Co-expression of *PLAU* (25.4%) and its cognate receptor *PLAUR* (75.8%) provided transcriptional evidence for the predicted autocrine loop. These MDSC-associated transcripts were barely detected in pDCs, confirming the compartmentalization of signaling components.

Chord diagrams revealed that MIF signals converging on GZMB+ pDCs originated from diverse sources, including malignant epithelial cells, fibroblasts, and mast cells (Figure 2E, left). A similar pattern characterized the TGF-β network, though these incoming signals were immune-derived (mast cells, T and B lymphocytes) (Supplementary Figure S3A). ISG+ PMN-MDSC-derived PLAU targeted the broader myeloid compartment and the MDSCs themselves (Figure 2E, right). The remaining MDSC-derived axes acted on distinct downstream targets (Supplementary Figure S3B): endothelial cells for VISFATIN and CXCL, stromal pericytes for OSM, and the MDSC population itself for the IL-1 autocrine loop.

### 3.3 Spatial transcriptomics mapping validates the compartmentalized architecture and physical interactome of the dual-core subsets

Exhibiting the highest Golden Index under the SIDI framework (Supplementary Table S2), slice PT13 was prioritized for spatial mapping. By aligning a transcriptomic Niche Index with Visium coordinates, we reconstructed the tissue architecture. This generated a continuous spatial gradient (Figure 3A) that partitioned the microenvironment into three histological domains: malignant tumor nests, desmoplastic stroma, and the invasive tumor-stroma frontline (Δ*Score*=0).

**Figure 3.**
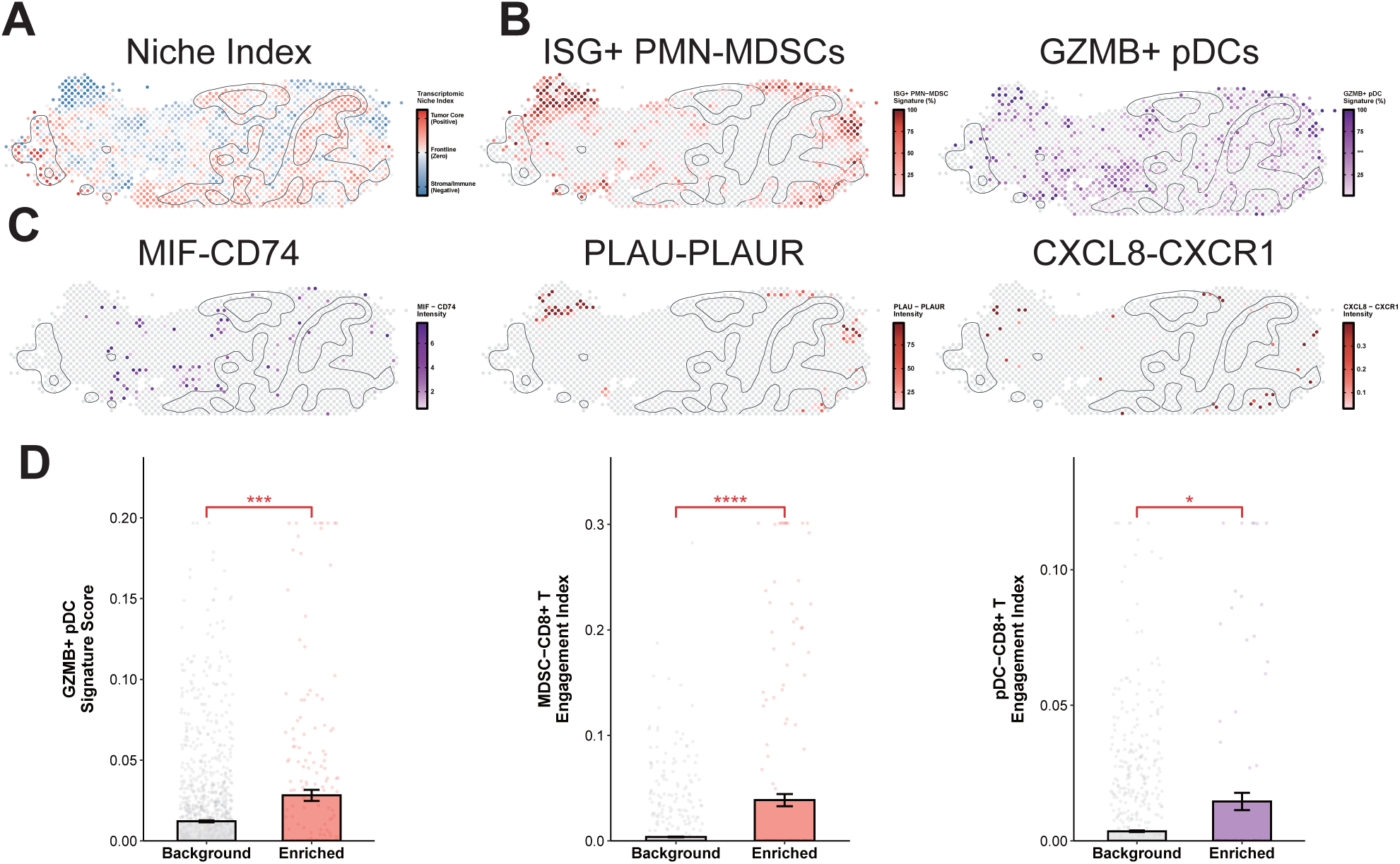
Spatial transcriptomic mapping validates the compartmentalized architecture and physical interactome of the dual-core subsets. **(A)** Spatial mapping of the transcriptomic Niche Index for the OSCC slice PT13. The color gradient (ΔScore) delineates the tumor core (red), stroma (blue), and transitional invasive margin (white). **(B)** Spatial signature mapping of ISG+ PMN-MDSCs (left) and GZMB+ pDCs (right). Spot colors indicate relative signature scores (0-100%). Dark-grey contours demarcate the tumor boundary. **(C)** Spatial mapping of core intercellular signaling axes. The color gradient reflects the geometric interaction intensity. Panels display the incoming MIF-CD74 (left) alongside MDSC-derived PLAU-PLAUR (middle) and CXCL8-CXCR1 (right) interactions. **(D)** Spatial niche enrichment and geometric engagement analysis. Bar plots (mean ± SEM) with individual data points compare GZMB+ pDC abundance in MDSC-enriched niches versus background stroma (left). The middle and right panels quantify localized interactions with CD8+ T cells using the geometric Engagement Index (*E_i_*). Statistical significance was determined by the Wilcoxon rank-sum test (*P < 0.05, ***P < 0.001, ****P < 0.0001).

Spatial signature mapping revealed that ISG+ PMN-MDSCs aggregated outside the tumor contour to form a boundary along the invasive margin (Figure 3B), while GZMB+ pDCs penetrated into the solid tumor core. This spatial segregation places the pDCs in direct physical proximity to the malignant epithelial compartment, whereas the MDSCs are positioned to interact with the surrounding stroma. Consistent with this spatial segregation, MIF-CD74 signaling hotzones spanned the stroma and breached the solid tumor contour. The MDSC-associated pathways maintained strict extra-tumoral confinement: PLAU-PLAUR interactions localized entirely within the stroma without penetrating the tumor nests, and CXCL8-CXCR1 signaling accumulated at the invasive margin (Figure 3C).

Quantitative niche analysis corroborated these distributions (Figure 3D). Although a subset of pDCs penetrated the tumor core, MDSC-enriched niches at the invasive margin harbored a significantly higher abundance of GZMB+ pDCs compared to the background stroma (P = 0.0001), alongside co-localization with CD8+ T cells (P < 0.0001). This spatial interactome was conserved across six independent OSCC cohorts (Supplementary Table S3): pDCs and MDSCs co-localized with CD8+ T cells in 100% (6/6) and 83% (5/6) of the datasets, respectively, while direct pDC-MDSC proximity occurred in 67% (4/6) of cases.

### 3.4 In silico ablation reveals the structural dependency of the global TME network on the dual-core subsets

We performed *in silico* perturbation to evaluate the fragility of the TME network (Figure 4A, top). Relative to the unperturbed baseline, computational ablation of ISG+ PMN-MDSCs and GZMB+ pDCs collapsed the global communication flux, reducing total intercellular interaction probabilities to 42% and 35%, respectively (Figure 4A, bottom). Ablation of ISG+ PMN-MDSCs broadly disrupted the ANNEXIN, PLAU, IL1, VISFATIN, and CXCL axes (Figure 4B), while GZMB+ pDC depletion drove a restricted degradation of the MIF signaling axis (Figure 4C).

**Figure 4.**
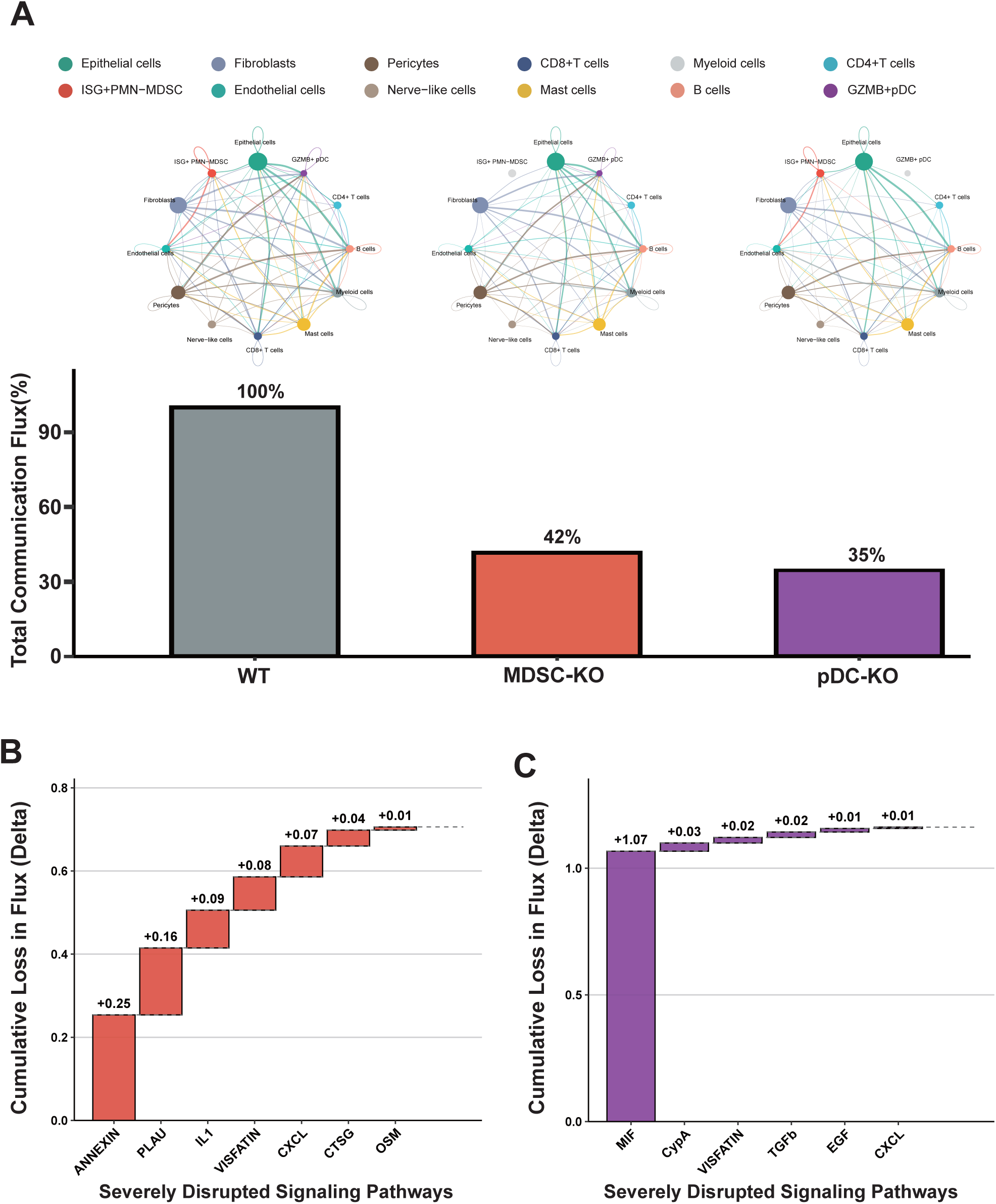
In silico ablation reveals the structural dependency of the global TME network on ISG+ PMN-MDSCs and GZMB+ pDCs. (A) Circle plots of the OSCC intercellular communication network at baseline (left) and after ablation of ISG+ PMN-MDSCs (middle) or GZMB+ pDCs (right). Nodes represent cell lineages, with edge widths reflecting signaling weights. The bottom panel quantifies total communication flux. (B-C) Waterfall plots of absolute flux loss (ΔFlux) across signaling pathways after ablation of ISG+ PMN-MDSCs (B) and GZMB+ pDCs (C). Pathways are ordered by flux loss.

### 3.5 Construction and external validation of the 5-gene prognostic signature

While the initial 7-gene panel characterized intracellular mitoxyperilysis, we leveraged downstream intercellular communication networks to perform LASSO Cox regression, and isolated a 5-gene signature (*CTSG*, *NAMPT*, *AREG*, *PLAU*, and *CXCL8*) (Figure 5A; Supplementary Table S4). Within the TCGA-OSCC cohort, high-risk patients exhibited significantly shorter overall survival (OS) (P < 0.001, Figure 5B), with risk score plots and heatmaps confirming distinct gene expression patterns between the risk groups (Supplementary Figure S4). Multivariate Cox regression validated this signature as an independent prognostic factor (HR = 2.18, 95% CI: 1.37-3.47, P = 0.001; Figure 5C; Supplementary Table S5). Beyond the risk score, advanced clinical stage remained the only independent predictor of poor survival (HR = 2.25, 95% CI: 1.18-4.28, P = 0.014); age and gender lacked statistical significance. Time-dependent ROC analysis confirmed the predictive accuracy of the signature, yielding 1-, 3-, and 5-year AUCs of 0.640, 0.637, and 0.630, respectively (Figure 5D).

**Figure 5.**
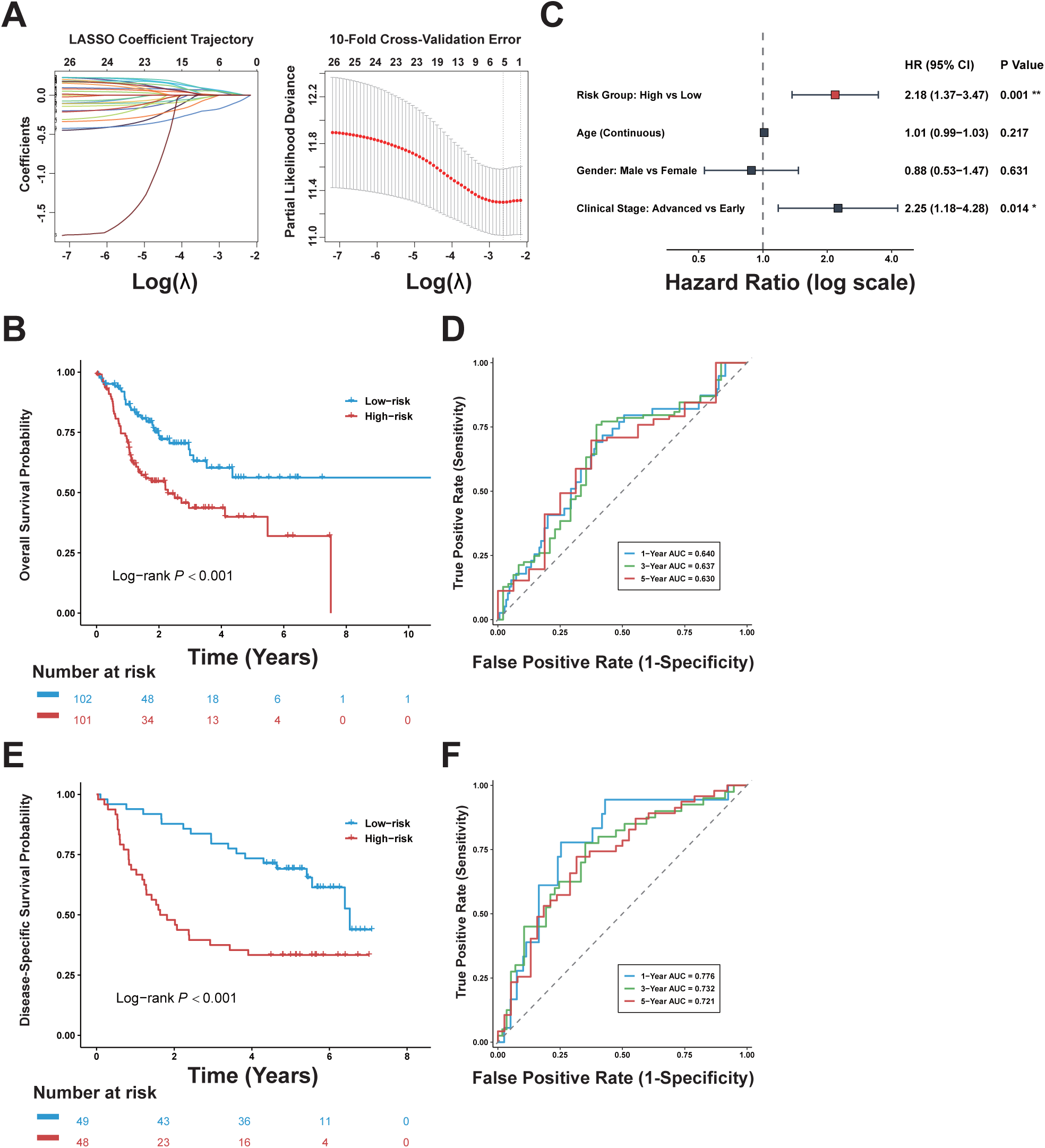
Construction and multi-cohort validation of the 5-gene prognostic signature. (A) LASSO coefficient profiles and 10-fold cross-validation curves. (B) Kaplan-Meier overall survival (OS) curves in the TCGA-OSCC cohort. (C) Forest plot of multivariate Cox regression analysis. (D) Time-dependent ROC curves at 1, 3, and 5 years in the TCGA-OSCC cohort. (E) Kaplan-Meier disease-specific survival (DSS) curves in the GSE41613 cohort. (F) Time-dependent ROC curves at 1, 3, and 5 years in the GSE41613 cohort.

For external validation in the GSE41613 cohort, high-risk patients exhibited inferior disease-specific survival (DSS) (P = 0.002; Figure 5E), with the signature demonstrating strong discriminative capacity for tumor-specific mortality (1-, 3-, and 5-year AUCs of 0.776, 0.732, and 0.721, respectively; Figure 5F).

### 3.6 The 5-gene signature identifies an immune desert phenotype driven by spatial exclusion

Immunological profiling of the TME revealed that the continuous risk score inversely correlated with infiltrating immune populations (Figure 6A), driving a pan-immune desert phenotype that encompassed both effector lineages (e.g., CD8+ T cells, NK cells, B cells) and immunosuppressive compartments (e.g., MDSCs, Tregs).

**Figure 6.**
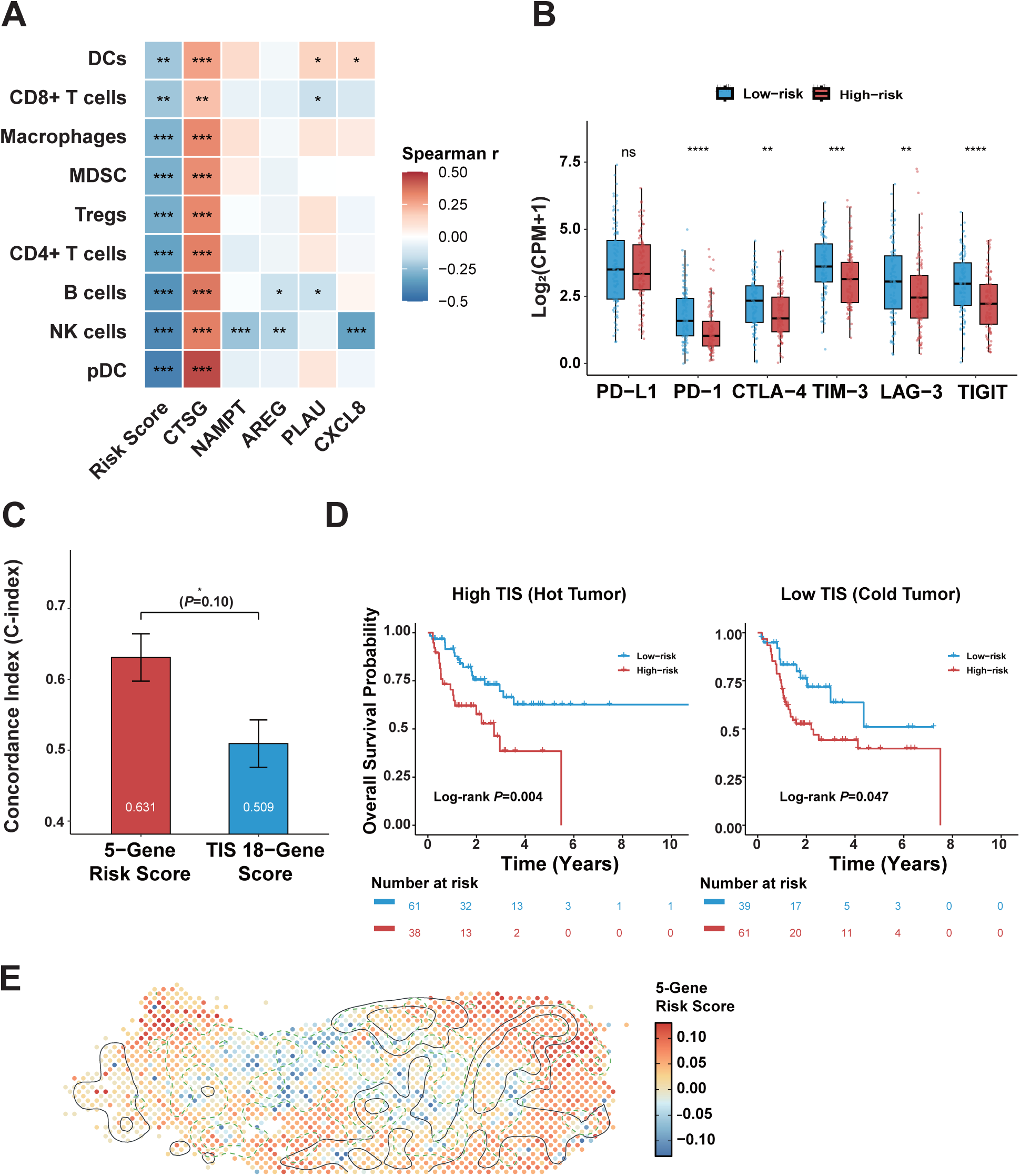
Characterization of the tumor immune microenvironment and immunotherapeutic potential. **(A)** Heatmap of Spearman correlations between the 5-gene risk score and immune cell infiltration. **(B)** Bar plot comparing the prognostic Concordance index (C-index) of the 5-gene risk score and the 18-gene Tumor Inflammation Signature (TIS). **(C)** Kaplan-Meier overall survival (OS) curves stratified by risk score within High TIS and Low TIS subgroups. **(D)** Boxplots of core immune checkpoint expression (PD-L1, PD-1, CTLA-4, TIM-3, LAG-3, and TIGIT). Statistical significance was determined by the Wilcoxon rank-sum test. **(E)** Spatial mapping of the 5-gene risk score and cytotoxic immune niches in slice PT13. Spot colors indicate the continuous risk score. Black and dashed green contours delineate the tumor core and cytotoxic niches, respectively. (ns, not significant; *P < 0.05, **P < 0.01, ***P < 0.001, ****P < 0.0001).

The expression of key co-inhibitory receptors (PD-1, CTLA4, TIM-3, LAG3, and TIGIT) was downregulated in high-risk patients (Figure 6B), whereas PD-L1 (CD274) levels remained unchanged. Comparison with the established 18-gene Tumor Inflammation Signature (TIS) confirmed the higher prognostic accuracy of our model (C-index: 0.631 vs. 0.509, P = 0.01; Figure 6C; Supplementary Table S6)**, and** the 5-gene signature successfully stratified patients within both the high and low TIS subgroups (Figure 6D).

In spatial transcriptomic (ST) mapping, high-risk regions localized to the extra-tumoral stroma, physically isolated from the cytotoxic immune core (Figure 6E). Conversely, low-to-moderate risk domains co-localized with active immune niches. This physical isolation mirrors the immune depletion phenotype observed in our bulk cohorts.

### 3.7 Construction and evaluation of a clinical prognostic nomogram

We constructed a prognostic nomogram integrating the 5-gene risk score, patient age, and clinical stage, yielding a C-index of 0.6529 for overall survival (Figure 7A). Calibration curves demonstrated strong concordance between predicted and observed survival probabilities at 1, 3, and 5 years (Figure 7B).

**Figure 7.**
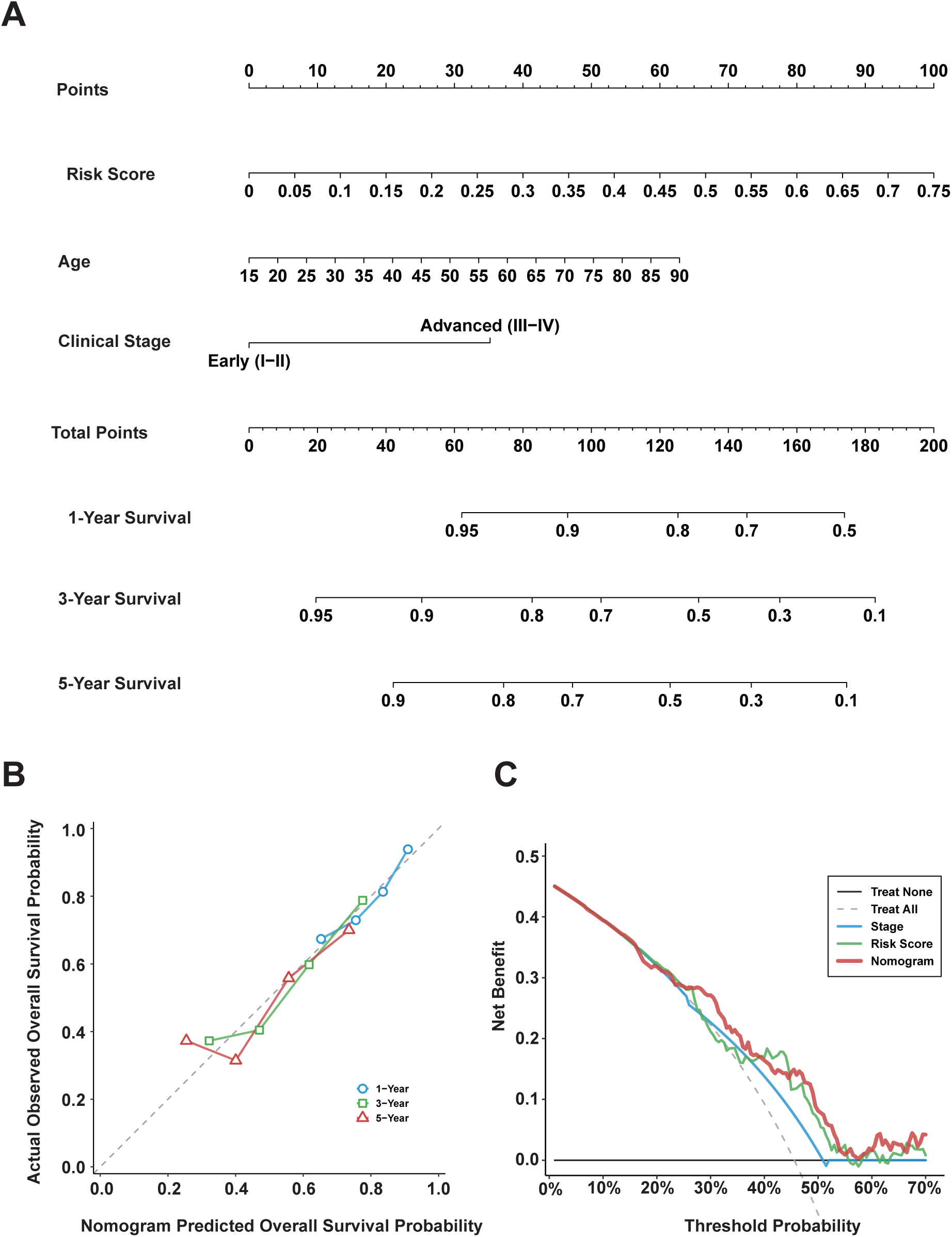
Construction and clinical utility of the integrated prognostic nomogram. (A) Nomogram predicting 1-, 3-, and 5-year overall survival (OS) based on the 5-gene risk score, age, and TNM stage. (B) Calibration curves for 1-, 3-, and 5-year OS predictions. Points represent bootstrap-corrected estimates (1000 resamples). (C) Decision curve analysis (DCA) comparing the net benefit of the nomogram, TNM stage, and 5-gene risk score at 3 years.

Decision curve analysis (DCA) evaluated the net clinical benefit of the predictive models (Figure 7C). Within the 20% to 50% threshold probability range, both the 5-gene risk score and the integrated nomogram provided greater net benefit than clinical stage alone. Although the nomogram yielded the highest benefit across most thresholds, it experienced a marginal dip between 16% and 20%. Notably, within the 40% to 45% interval, the standalone risk score exhibited locally superior performance, outperforming both the clinical stage and the integrated nomogram.

## 4. Discussion

The spatial architecture of the tumor microenvironment (TME) is a major determinant of immune evasion and therapeutic resistance in oral squamous cell carcinoma (OSCC) ^15^ ^33^ ^34^. Our findings provide spatial evidence that immune exclusion in OSCC is a barrier driven by localized mitoxyperilysis. Rather than reflecting generalized immune exhaustion, the cooperative interaction between ISG+ PMN-MDSCs and GZMB+ pDCs highlights a structured immunosuppressive niche. By translating this complex spatial phenotype into a 5-gene signature, we provide a reliable molecular surrogate for this barricade. The nomogram successfully integrates these spatial TME features with standard clinical staging.

Our spatial observations support a two-tiered metabolic-immune cascade driving this immunosuppressive network. Tumor-intrinsic metabolic stress—specifically, elevated mitochondrial reactive oxygen species (ROS)—stimulates the secretion of chemokines (e.g., CXCL2, G-CSF) that recruit myeloid-derived suppressor cells (MDSCs) and plasmacytoid dendritic cells (pDCs) to the invasive margin ^35^. Our OSCC spatial mapping reveals that upon converging at this metabolically hostile niche, specific subpopulations of these recruited MDSCs and pDCs undergo localized mitoxyperilysis. Characterized as a lytic cell death mechanism driven by prolonged mitochondria-plasma membrane contact, mitoxyperilysis causes the chronic extracellular release of ROS and damage-associated molecular patterns (DAMPs) such as HMGB1 ^13^ ^36–38^. Chronic exposure to these factors triggers a maladaptive Type I interferon (IFN) response in surrounding myeloid populations ^39^ ^40^,driving the emergence of the suppressive ISG+ PMN-MDSCs and GZMB+ pDCs subsets.

Our study reveals that localized mitoxyperilysis drives ISG+ PMN-MDSCs and GZMB+ pDCs to accumulate at the invasive margin, where they cooperate to form a stromal barrier. Spatial Niche Enrichment and Engagement Index analyses across multiple cohorts confirm this colocalization. While traditionally viewed as independent drivers of immune tolerance^39^ ^41^, our models reveal a complementary relationship. Rather than engaging in direct cell-to-cell cross-talk, these populations mediate immune exclusion through parallel signaling pathways. ISG+ PMN-MDSCs act as the primary signal emitters ^42^ ^43^, relying on both autocrine maintenance and paracrine remodeling. An IL-1 autocrine loop sustains their suppressive state ^44^, while these MDSCs concurrently emit signals (e.g., VISFATIN, OSM, PLAU, CXCL) targeting adjacent endothelial cells and pericytes. This active stromal remodeling restricts T cell infiltration.

GZMB+ pDCs function as receivers for the MIF-CD74 axis ^45^. While MDSCs build the physical stromal barrier, this MIF-CD74 axis connects the immune margin directly to the malignant nests. These MIF signals come from multiple sources—including malignant epithelial cells, fibroblasts, and mast cells—indicating that the regulatory GZMB+ phenotype is not driven by the tumor alone. By secreting GZMB, these pDCs degrade the TCR ζ-chain to suppress T-cell expansion ^46^ ^47^. Computationally ablating either the MDSC or pDC population causes a severe drop in overall signaling (58% and 65%, respectively), revealing that both subsets are required to maintain immune suppression in the TME ^48^.

TNM staging remains an essential reference for OSCC treatment and prognosis, yet it overlooks TME heterogeneity ^49^ ^50^. While recent single-cell and bulk analyses attempt to identify markers reflecting this heterogeneity ^51^ ^52^, they inherently ignore spatial context. By integrating single-cell and spatial transcriptomics, we screened the communication networks of the two mitoxyperilysis-related subpopulations to identify an independent prognostic gene signature. Although our initial 7-gene panel reflects the intracellular mitoxyperilysis state, monitoring downstream cell-to-cell communication minimizes the transcriptomic noise of generic markers and provides a more stable readout for routine prognosis. Beyond pure statistical significance, this signature directly mirrors the MDSC-pDC communication network, serving as a spatial proxy for the *in situ* status of these mitoxyperilysis-driven subpopulations.

Our DCA highlights the complementary roles of molecular and anatomical profiling. For general clinical decision-making (thresholds >20%), combining the TME landscape with clinical staging via the nomogram maximizes patient benefit. However, at high-risk thresholds (40%–45%), the standalone risk score outperforms the integrated nomogram. This reveals that for the most critical patients, the MDSC-pDC barrier dictates outcomes more than traditional tumor stage, meaning that adding clinical stage actually reduces predictive accuracy, thereby underscoring the signature’s independent power for identifying high-risk individuals.

Therapeutically, this signature identifies anatomically early-stage patients who already harbor an immune-desert microenvironment. These high-risk tumors exhibit global T-cell depletion alongside downregulated immune checkpoints (e.g., PD-1, CTLA-4, HAVCR2). Notably, PD-L1 expression remained unchanged. Because these patients lack the effector cells and targets required for conventional immune checkpoint blockade (ICB), reliance on PD-L1 expression alone may mislead the use of anti-PD-1/PD-L1 monotherapies, yielding limited clinical benefit ^7^ ^53^. This spatial exclusion also creates a blind spot for bulk inflammatory metrics like the 18-gene TIS^26^. A high TIS score means little if effector cells are physically trapped outside the tumor^54^ ^55^. By quantifying this stromal barricade, the 5-gene signature complements TIS and stratifies risk even within highly inflamed subgroups. Thees findings provide a biological rationale for exploring combination strategies—such as stroma-targeting or myeloid-modulating agents—to remodel this physical barrier before attempting T-cell activation^56–58^.

Several limitations of this study should be acknowledged. Methodologically, our prognostic models rely on retrospective cohorts and require prospective, multicenter validation. Because standard spatial transcriptomics lacks true single-cell resolution, future sub-cellular spatial profiling and multiplexed proteomics ^59^ are needed to map these exact cellular interfaces and confirm the mitoxyperilysis-driven metabolic states at the protein level. Clinically, transcriptomic PD-L1 (*CD274*) expression may not fully align with the Combined Positive Score (CPS) determined by standard immunohistochemistry^4^ ^60^.

## Conclusion

Our study defines an immunosuppressive network in OSCC driven by mitoxyperilysis. We show that myeloid accumulation at the tumor margin creates a physical and metabolic barrier that restricts T-cell infiltration. Translating these spatial insights into a validated 5-gene nomogram provides a practical tool for patient risk stratification. Targeting this myeloid-stromal axis offers a strategy to overcome immune exclusion, and developing a multiplexed immunohistochemistry (mIHC) panel based on our signature represents the essential next step for routine clinical application.

## Data Availability

Publicly available datasets analyzed in this study are accessible via GEO (GSE310797, GSE41613) and TCGA (TCGA-HNSC). Derived data and modeling coefficients are provided in the Supplementary Information. Custom R scripts used for all computational analyses are publicly available via GitHub at https://github.com/zoubo9465/OSCC-Mitoxyperilysis-Analysis. Additional information required to reanalyze the data is available from the corresponding author upon request.

## Declarations

### Author Contributions

**B.Z.** conceptualized and supervised the study. **J.L.** and **D.Z.** performed the bioinformatics analysis, spatial transcriptomic mapping, and drafted the manuscript. **Z.M.** contributed to data curation, methodology validation, and figure preparation. **B.Z.**, and **Z.M.** critically revised the manuscript. All authors have read and approved the final submission.

### Funding

This work was supported by the General Program of the Scientific Research Development Fund of the Affiliated Hospital of Shandong Second Medical University (Grant No. 2025FYM128) and the Natural Science Foundation of Liaocheng People’s Hospital (Grant No. LYQN201916).

### Declaration of Interests

The authors declare no competing interests.

### Ethics Approval and Consent to Participate

All patient data analyzed in this study were obtained from publicly available databases (TCGA, GEO). The original studies obtained ethical approval and patient consent. Therefore, additional local ethical approval was waived.

## Notes

### Competing Interest Statement

The authors have declared no competing interest.

### Author Declarations

The study used ONLY openly available human data that were openly accessible before the initiation of the study. The original source data can be located and downloaded at the following public repositories: 1. Gene Expression Omnibus (GEO): Accession numbers GSE310797 (https://www.ncbi.nlm.nih.gov/geo/query/acc.cgi?acc=GSE310797) and GSE41613 (https://www.ncbi.nlm.nih.gov/geo/query/acc.cgi?acc=GSE41613). 2. The Cancer Genome Atlas (TCGA): TCGA-HNSC dataset, accessible via the NIH GDC Data Portal (https://portal.gdc.cancer.gov/projects/TCGA-HNSC).

