## Supplementary figures and images for "A mitoxyperilysis-derived spatial signature predicts immune exclusion and survival in oral squamous cell carcinoma"

### Supplementary Figures

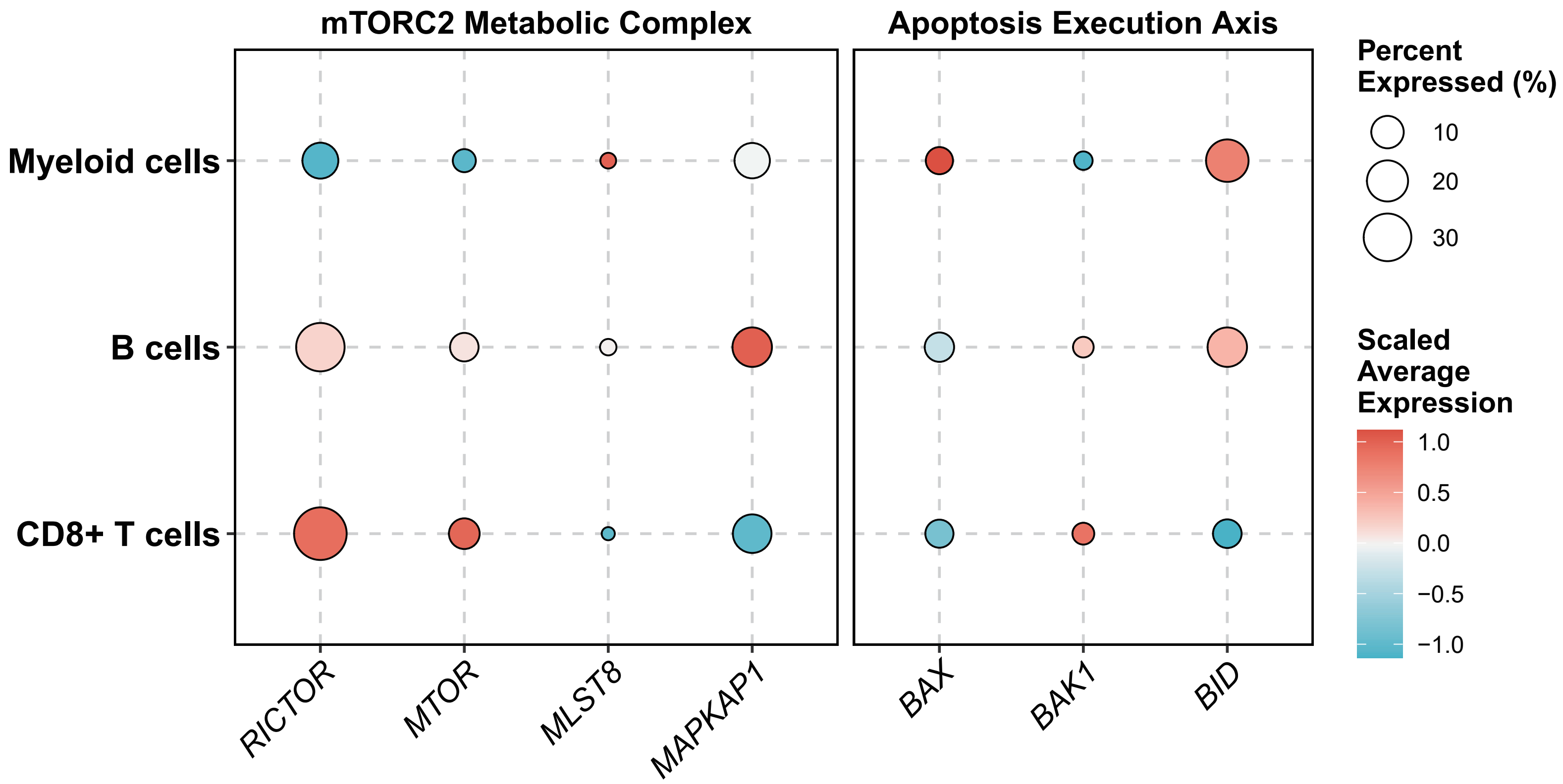

### Supplementary Figures

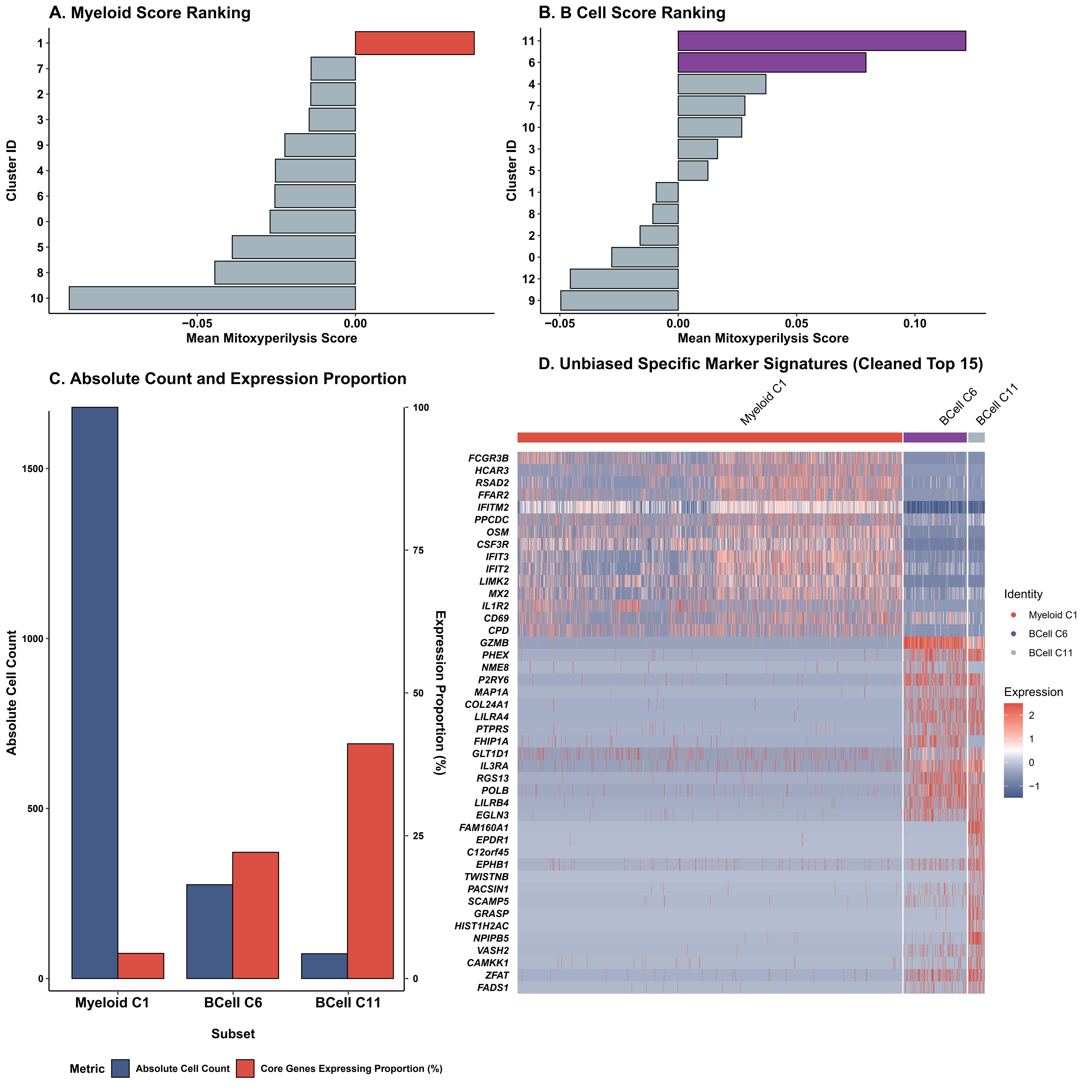

### Supplementary Figures

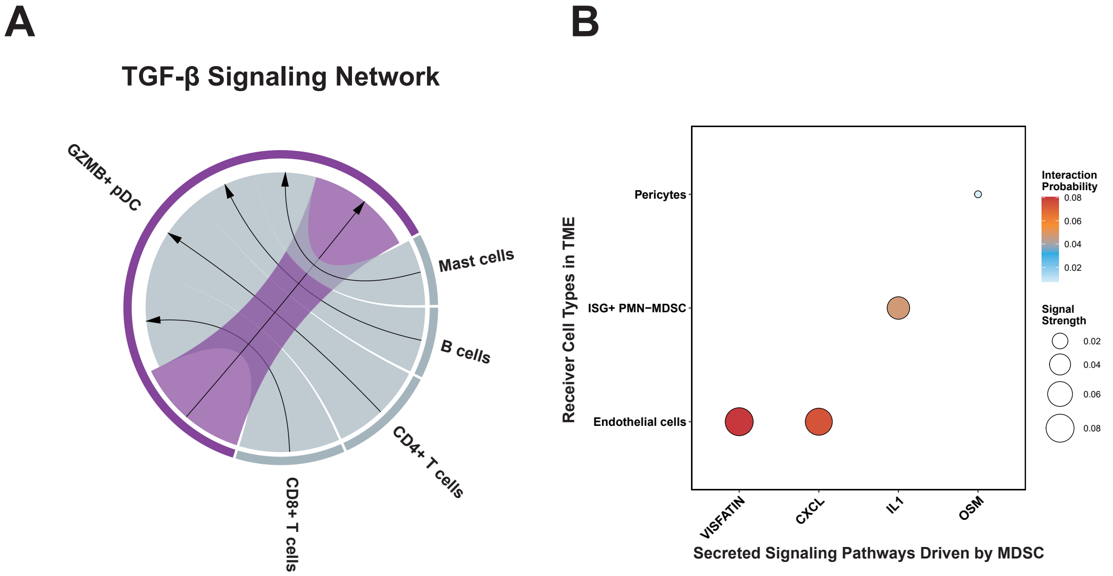

### Supplementary Figures

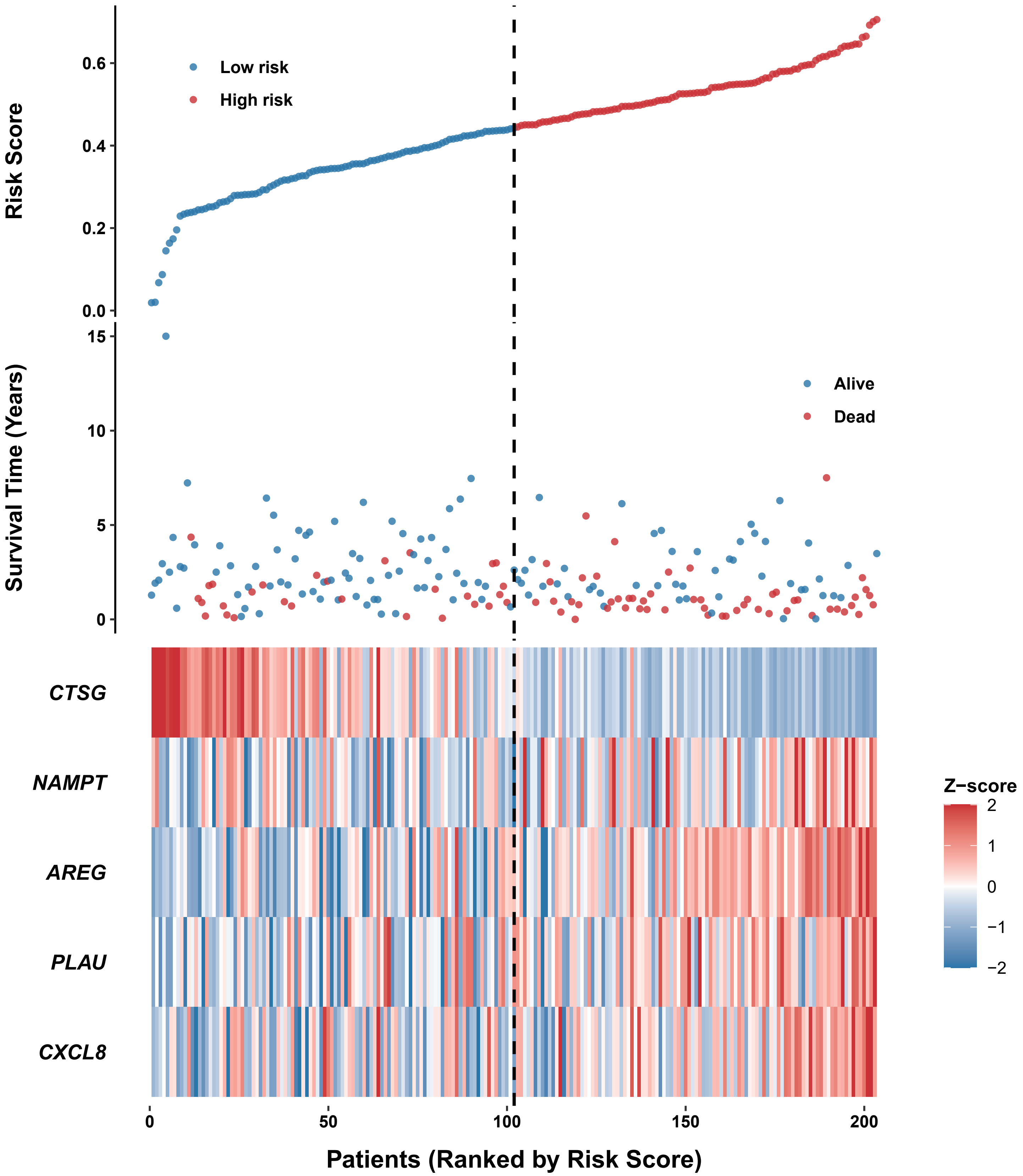
